# Preoperative Intensification Treatment in Patients with Rectal Adenocarcinoma – A Standardization of Clinical Practice

**DOI:** 10.1101/2023.01.09.23284351

**Authors:** Luísa Leal-Costa, Diana Silva, Carlota Baptista, Rita Bizarro, Madalena Machete, Pedro Simões, Ana Faria, José Alberto Teixeira

## Abstract

Neoadjuvant radiotherapy with concurrent fluoropyrimidines followed by surgery is considered the standard-of-care in locally advanced rectal cancer (LARC). Neoadjuvant chemo-radiotherapy (CRT) is associated with a pathological complete response (pCR) rate of 11-18%. Intensification of neoadjuvant treatment can lead to higher rates of tumor resectability and pCR, although the best therapeutic sequence is unknown.

This is a prospective, single arm study which aims to standardize institutional clinical practice in accordance with international recommendations. Patients with LARC received neoadjuvant intensive treatment with concurrent CRT followed by 12 weeks of consolidation chemotherapy (ChT) with CAPOX or mFOLFOX6. Clinical response was accessed by magnetic resonance imaging (MRI) at 10 weeks, 14 weeks, and 18 weeks. If complete clinical response (cCR) was obtained, a *watch-and-wait* (W&W) strategy was offered; otherwise, surgery was performed. The primary endpoint was to evaluate clinical response rate after 10 weeks of intensification treatment, and its comparison with historical data. Key secondary endpoints included clinical response rate at 14 and 18 weeks, rate of patients who enter W&W strategy, rate of pCR in patients who undergo surgery, recurrence free survival (RFS), overall survival (OS), and rate of adverse effects. We present efficacy and safety preliminary results one year from the start of the study.

Six patients with LARC were included. Three had a tumour in the low-rectum, two in middle-rectum and one in high-rectum. At initial magnetic resonance imaging (MRI), two had cT2, three had cT3 and one had cT4; three had cN1 and one cN2; two patients had extramural venous invasion (EMVI+) and one had mesorectal fascia involvement (MRF+).

After CRT, all patients started consolidation ChT. The most common grade ≥3 AEs were neutropenia, nausea, and diarrhoea. There were no dose-limiting toxicities and all patients completed treatment. MRI at 10 weeks showed a tumour regression grade (TRG) 2 in three patients, TRG3 in two patients and TRG4 in one patient. Two patients are in the W&W surveillance protocol; one is awaiting MRI at 18 weeks; another patient awaits surgery; and of the two patients already underwent surgery, one had a ypT3N1 and the other had complete pathological response (pCR). To this date there are no recurrence or death events.

These preliminary results suggests tolerability and feasibility of a neoadjuvant intensification treatment in patients with LARC.

**Categories:** rectal adenocarcinoma, oncology

## Introduction

Rectal tumours include those arising up to 15cm from the anal margin (0-5cm lower-, 5-10cm middle-, and 10-15cm upper-rectum). Magnetic resonance imaging (MRI) is the preferred imaging modality for locoregional staging and risk stratification. Total mesorectal excision (TME) is the standard surgical option. Due to the anatomical proximity to other organs, surgical morbidity and mortality rates are not negligible. In addition to a permanent stoma, many patients are left with sequelae of faecal/urinary incontinence and/or sexual dysfunction, with a great impact in quality of life.

European Society of Medical Oncology (ESMO) (1) and National Comprehensive Cancer Network (NCCN) (2) guidelines suggest intensifying treatment in higher risk tumours (positive extramural venous invasion (EMVI) or mesorectal fascia involvement (MRF), ≥cT3c, N+, elevator ani muscles involvement). This strategy, commonly known as Total Neoadjuvant Therapy (TNT), has the potential to increase response rates, including complete response (pCR), which stands at 11-18% with standard chemo-radiotherapy CRT [3–6]. TNT can also improve compliance and decrease recurrence rates (3). Some studies demonstrate a correlation between complete response rates and survival (7).

Recent data have shown that, after TNT, patients with clinical complete response (cCR) can be actively monitored without radical resection, through a strict follow-up protocol of proctologic, endoscopic and MRI assessments [8–13]. This approach is called *watch-and-wait* (W&W) and has been pursued by several international groups, to allow for organ preservation without compromising survival outcomes [14–17]. Currently, our institution has a prospective study in this area, which increases the need of standardizing neoadjuvant intensification therapy.

In 2020, two multicentre phase III studies, RAPIDO (18) and PRODIGE-23 (19), studied the potential benefit of TNT strategies in patients with rectal adenocarcinoma. Both selected patients ≥18 years, with locally advanced rectal cancers (LARC): cT4 or cT3 with high-risk criteria (EMVI+, MRF+, N2 or suspicious lateral lymph nodes).

RAPIDO trial (18) compared standard CRT with an experimental arm of short course radiotherapy (RT) regimen followed by 18-week period of consolidation FOLFOX4/CAPOX, both arms followed by TME. Adjuvant chemotherapy (ChT) was optional in the control arm. The pCR rate was 27.7% vs 13.8% (OR 2.40 (1.70-3.39); p<0.001) in the experimental and standard arms, respectively, and a lower rate of disease recurrence (23.7% vs 30.4%; p=0.019) was found in high-risk LARC patients treated with the TNT strategy compared to conventional CRT.

PRODIGE-23 trial (19) compared isolated standard CRT to a control arm of induction ChT triplet with FOLFIRINOX for six cycles followed by neoadjuvant CRT, both arms followed by TME. After surgery, patients completed adjuvant treatment with six cycles of mFOLFOX6 or four cycles of capecitabine, depending on choice of centre. The pCR rate was 27.5% in the experimental arm vs 11.7% in the standard arm (p<0.001). Neoadjuvant FOLFIRINOX followed by CRT significantly increased recurrence-free survival (RFS) (76% vs 69%, p=0.034) and metastasis-free survival (MFS) (79% vs 72%, p=0.017) with no significant increase in toxicity.

Two more trials studying the benefit of TNT were published in 2022.

STELLAR (20) was a noninferiority trial which included patients with distal or middle-third, cT3-4 and/or N+. The experimental arm was similar to RAPIDO (18) and tested SCRT followed by four cycles of CAPOX; the standard arm was treated with CRT concurrent with capecitabine. TME was performed 6-8 weeks after neoadjuvant treatment. After surgery, patients completed two additional cycles of CAPOX in the experimental group and six cycles in the CRT group. The 3-year disease free survival (DFS) was 64.5% and 62.3% in TNT and CRT groups, respectively (p<0.001 for noninferiority). Although there was no significant difference in MFS or locoregional recurrence, the TNT group had better 3-year overall survival (OS) than the CRT group (86.5% vs 75.1%, p= 0.033).

OPRA trial (21) randomized patients with clinical stage II and III distal rectal cancer to 4 months of mFOLFOX6 or CAPOX before (induction group (INCT-CRT)) or after (consolidation group (CRT-CNCT)) standard CRT. Patients were re-staged at 8-12 weeks after completion of TNT with rectal examination, sigmoidoscopy, and MRI. Those who achieved cCR or near-complete response were offered a W&W strategy; the others underwent TME. This study concluded that the W&W strategy in patients who achieve a cCR after TNT allows for organ preservation in a high percentage of patients, higher in consolidation arm (41% vs 53%). No differences were found between groups in local RSF, MFS, or OS.

These trials demonstrate that TNT is associated with higher response and organ preservation rates, particularly consolidation ChT. Furthermore, a potential survival benefit should not be ignored for high tumours of middle-and upper-rectum (included in RAPIDO (18) and PRODIGE-23 (19) trials).

## Materials & Methods

### Study design

This is a prospective single-centre cohort study. It was designed to recruit 16 patients per year until 1^st^ June 2026. A preliminary assessment was intended one year from the start of the study to evaluate the efficacy and safety of this strategy. Final data processing and final publication is planned between June and August 2026.

The primary endpoint was to evaluate the cCR rate at 10 weeks of neoadjuvant intensification treatment and compare it with historical data.

cCR rate was defined as the presence of all of the following criteria: MRI without evidence of residual lesion (the presence of fibrosis or oedema is not considered) and no lymph nodes with suspicion criteria – tumour regression grade 1 (TRG1); absence of a palpable lesion on digital rectal examination (a subtle decrease in elasticity in the scar area is acceptable); and a rectosigmoidoscopy without residual lesion (the presence of a white scar or telangiectasia is acceptable). Good response was defined as the presence of all of the following criteria: MRI with evidence of a significant lesion reduction – TRG2; absence of a palpable lesion on digital rectal examination (a small, smooth superficial lesion or a subtle decrease in elasticity in the scar area is acceptable); and a rectosigmoidoscopy with either no evidence of residual lesion (the presence of a white scar or telangiectasias is acceptable) or with only minor changes (slight mucosal irregularity, flat and superficial ulcer <1cm, and/or mucosal erythema). A poor/non-response was considered in patients who did not meet the criteria for cCR (TRG3-4). pCR was defined as ypT0/is ypN0 in patients undergoing surgery.

The secondary endpoints were to evaluate cCR conversion rate at 14 weeks and 18 weeks of neoadjuvant intensification treatment, in patients who did not reach cCR at 10 weeks; to quantify the proportion of patients who enter a W&W strategy; to determine OS and RFS in patients undergoing neoadjuvant intensification treatment; to characterize adverse effects; and to study the correlation between cCR and pCR after neoadjuvant intensifying treatment in patients undergoing surgery.

OS was defined as the time from diagnosis to death from any cause. RFS was defined as the time from diagnosis to relapse documented by imaging or biopsy, or death from any cause.

### Patients

All patients with LARC discussed in a multidisciplinary gastrointestinal tumour board from 1^st^ June 2021 and who have given written informed consent were recruited.

Inclusion criteria were age ≥18 years; ability to understand the risks and sign informed consent before starting the protocol; histological confirmation of adenocarcinoma of the rectum with the following characteristics: lower rectal tumours not candidates for endoscopic resection or minimally invasive transanal surgery regardless of cTN, middle/high rectal tumours ≥cT3c/d and/or cN2 and/or MRF+ and/or EMVI+; staging without evidence of metastasis (cM0); and ability to comply with neoadjuvant ChT and RT treatments. Exclusion criteria were tumours that are candidates for direct surgery; synchronous neoplasms; prior pelvic RT; previous rectal surgery; contraindication for ChT; contraindication for MRI; and patients with second active neoplasm under investigation or treatment at the time of diagnosis.

### Procedures

All patients were staged by pelvic MRI and thoracoabdominopelvic CT scan. All patients considered to have a LARC diagnosis were selected from the multidisciplinary gastrointestinal tumour board.

The protocol flow diagram is shown in **Figure 1**.

**Figure 1.**
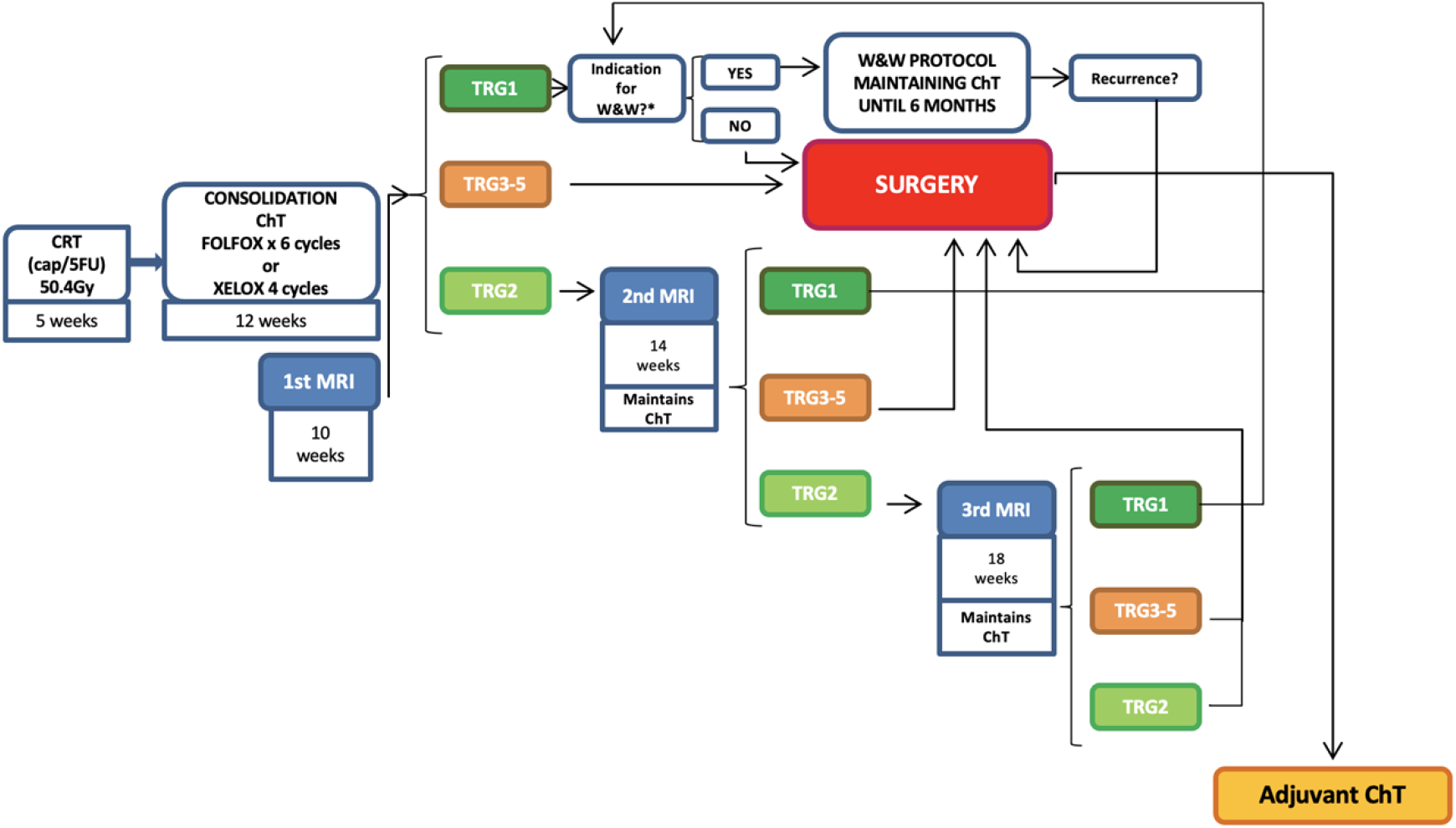
Protocol flow diagram. *middle/high rectal tumours were proposed for surgery even if TRG1.

Neoadjuvant treatment was started with concurrent CRT (50.4Gy during 5 weeks and 825mg/m^2^ concurrent oral (PO) capecitabine twice daily or fluorouracil (5FU) 225mg/m^2^/day continuous infusion over 5 days). This was followed by 12 weeks (ideally starting at week 6) of consolidation ChT with six cycles of mFOLFOX6 (oxaliplatin 85mg/m^2^ and leucovorin 400mg/m^2^ intravenous (IV) on day 1, followed by bolus 5FU 400mg/m^2^ IV and then continuous infusion at a dose of 2400mg/m^2^ over 46h every 14 days) or four cycles of CAPOX (capecitabine 1000mg/m^2^ PO twice daily on days 1-14 and oxaliplatin 130mg/m^2^ IV on day 1, every 21 days). In low rectum cT2N0 or cT3N0 tumours, which in theory do not benefit from adjuvant oxaliplatin by extrapolation from the MOSAIC trial (22), consolidation ChT was dependent on discussion with the patient and their consent, since the objective of the study is an increase in local response. The choice between CAPOX or mFOLFOX6 was per medical oncologist preference.

A second MRI was performed at 10 weeks (+/-7 days), after CRT completion for evaluation of treatment response, regardless of the ability to comply with ChT protocol. At this time, patients were on third cycle of mFOLFOX6 or second cycle of CAPOX.

For patients who met cCR criteria (TRG1) at 10 weeks, ChT was maintained up to a total of 6 months if cT3/cT4 and/or cN+ and/or EMVI+ at the time of diagnosis. A W&W strategy was then proposed. Surveillance was made with rectosigmoidoscopy, proctological examination and carcinoembryonic antigen (CEA) each 3 months in the first 2 years, and each 6 months in years 3 to 5; pelvic MRI each 3 months in the first year, each 6 months in the second year, and annually up to 5 years; and thoracoabdominal CT each 6 months in the first two years and annually up to 5 years.

Surgery with TME was proposed if evidence of local progression was found in any of the surveillance exams.

For patients with partial response criteria at 10 weeks, if MRI had TRG2 or nearly complete response, patients were reassessed with a second MRI at 14 weeks (fifth cycle of mFOLFOX6 or third cycle of CAPOX); if MRI had TRG3-4 or absence of partial response criteria, TME was offered.

For patients with partial response criteria at 14 weeks, if MRI had TRG2 or nearly complete response, patients were reassessed with a third MRI at 18 weeks (eighth cycle of mFOLFOX6 or fifth cycle of CAPOX); if MRI had TRG3-4 or absence of partial response criteria, TME was offered. All patients with partial response who repeated MRI at 14 and/or 18 weeks maintained ChT if they had not completed 6 months of treatment. A final reassessment was allowed at 18 weeks, after which, in the absence of complete response (TRG1), TME was planned.

### Data collection

Collected demographic, clinical and pathological variables were: gender; age at diagnosis; date of diagnosis; Eastern Cooperative Oncology Group Performance Status (ECOG PS); tumour location (low, middle or high rectum); TNM stage at diagnosis (American Joint Committee on Cancer system, eighth edition); presence of MRF invasion or EMVI; CEA level at diagnosis; CRT start and end dates; consolidation ChT start and end dates; ChT protocol and number of cycles; need of ChT dose reduction and/or defer (yes/no); ChT adverse effects (AEs); date of first MRI at 10 weeks after initiation of neoadjuvant treatment; CEA at the date of the first MRI; clinical restaging (ycTNM) and TRG at the date of the first MRI; date of second and third MRI with clinical restaging (ycTNM) and TRG if applicable; surgery (yes/no) and date; pathological staging (ypTNM); date and type of relapse (locoregional or distant); date of death; date of last follow-up.

Data were collected from electronic medical records (EMR). MRI were reviewed by radiologists with experience in colorectal cancer. Dates of death, when applicable, were retrieved from the Portuguese healthcare service National User Registry, which is updated daily. Data of patients lost to follow-up were considered up until the time they were last physically evaluated, after which they were censored for survival analysis.

### Statistical analysis

This is a single-arm prospective cohort study without a comparator arm. Based on the previous years of our institution, with 65 patients diagnosed with LARC in 4 years, we anticipated the inclusion of 16 patients per year.

Extrapolating from the pCR rates obtained in the RAPIDO (18) and PRODIGE-23 (19) trials (13% in the conventional neoadjuvant CRT arm and 28% in the experimental arm), we expected a cCR rate in our sample of 28%. To detect an increase in the rate of cCR from 13% to 28%, corresponding to an odds ratio (OR) of 2.6, 48 patients are required to achieve a power of 80% with a two-sided alpha level of 0.05. With the sample of 80 patients that we hope to recruit over the 5 years of the study, we will be able to reach a statistical power of 93.6% in the same comparison.

Continuous variables will be described as mean +/-standard deviation or median and interquartile range (IQR), depending on the distribution underlying the data. Categorical variables, such as cCR conversion rate at 10, 14 and 18 weeks, pCR rate and the proportion of patients who enter W&W strategy are presented as n (%). The mean of cCR will be compared with the values known in the literature through one sample t-test. The association between cCR and pCR after neoadjuvant intensification treatment in patients undergoing surgery will be performed through chi-square test.

Survival outcomes (OS and RFS) will be evaluated using the Kaplan-Meier estimator. A significance level of 5% will be considered in all statistical tests used.

Statistical analysis will be performed using IBM software-Statistical Package for Social Sciences® (SPSS) version 27.0. Sample calculation was done using STATA® (StataCorp LLC, College Station, TX, USA) software version 14.

### Ethical considerations

This study was approved by the Institutional Review Board/Independent Ethics Committee (RB/IEC) and designed according to Good Clinical Practice guidelines and the Declaration of Helsinki. All participants in this signed a written informed consent. Clinical data were treated with pseudonymization and kept accessible only to the primary investigator.

## Results

From august/2021 to august/2022, a total of 11 patients with LARC were considered eligible and gave written informed consent to participate in this study. Of these, only six had undergone treatment and the 10 week MRI.

The median age was 69.5 years old (range, 57-71), three patients were male and all had ECOG-PS 0. All tumours were adenocarcinomas; three were primarily located in low-rectum, two in middle-rectum and one in high-rectum.

At diagnosis, two patients were cT2, three cT3 and one cT4; two patients were cN0, three cN1 and one cN2; and two patients were EMVI+ and one was MRF+. The median of CEA at diagnosis was 2.6U/mL (range, 0-23.2).

In addition to CRT and prior to first MRI, four patients were treated with consolidation ChT with CAPOX and two patients with mFOLFOX6. The median time between the end of CRT and the first MRI was 9.8 weeks (range, 9.1-14.1), which corresponded to a mean of 3 (range, 2-4) cycles of CAPOX and 6 cycles of mFOLFOX6.

The most common grade ≥3 AEs were neutropenia (2), nausea (1), and diarrhoea (1). Other referred grade <3 AEs were anaemia (3), asthenia (2), thrombocytopenia (1), hepatotoxicity (1), and neurotoxicity (1). There were no dose-limiting toxicities and therefore no patient discontinued treatment; two patients had ChT dose reduction due to AEs (neutropenia and diarrhoea).

At the time of the first MRI (10 weeks after the end of CRT), the median CEA was 2.5U/mL (range, 1.1-3.7). The first MRI showed a TRG2 in three patients, TRG3 in two patients and TRG4 in one patient.

Patients with TRG2 repeated the second MRI at 14 weeks: two had a TRG1 and are in the W&W surveillance protocol; the third maintained TRG2, and is awaiting the third MRI at 18 weeks. Of the two patients with TRG3: one (middle-rectum, cT3N1, EMVI+/FMR-) has already been operated and the surgical specimen showed a ypT3N1 with ypTRG2, and the second (low-rectum, cT3bN1 EMVI-/MRF-) is currently awaiting surgery. The patient with TRG4 (high-rectum, cT4bN0 EMVI+/MRF+), underwent surgery and the surgical specimen showed ypT0N0 with ypTRG0. Until now, with a median time of follow-up of 8.5 months, no patients had tumour progression and no patients died.

## Discussion

Neoadjuvant CRT with concurrent fluoropyrimidines followed by surgery is considered the standard treatment in rectal cancer. However, the morbidity and mortality rates are not negligible, and sequelae of faecal/urinary incontinence and/or sexual dysfunction in addition to a permanent stoma have a great impact in quality of life. Randomized studies, retrospective analyses, and systematic reviews have been performed reporting excellent results in patients who achieve pCR after TNT, but there are no studies comparing the different treatment regimens.

Our study standardizes a TNT approach in patients with LARC and evaluates its cCR rates. Patients who achieved cCR or near-complete response were offered a W&W strategy and patients with incomplete response underwent TME. Two patients (33%) with a near complete response on first assessment at 10 weeks had a complete response after a 4 week delay and were candidates for organ preservation, which is consistent with data from clinical trials and suggests delaying surgery in good responders is feasible and effective.

Despite having a 33% organ preservation rate, one patient who underwent TME had a pCR, which amounts to 50% of both clinical and pathological complete responders within our study population. The chosen strategy of long course conventional CRT followed by consolidation ChT, as suggested by the OPRA trial (21), might lead to higher response rates and should be the preferred regimen for organ preservation. TNT regimens including SCRT, such as the RAPIDO (18) and STELLAR (20), might be more appropriate for ≤T3 high risk tumours for which survival benefit is the goal.

To date, it is not possible to state impact on OS and RFS in our population compared to historical data.

## Conclusions

In this preliminary analysis one year after from the start of the study, results suggest tolerability and feasibility of a neoadjuvant intensification treatment in patients with LARC. These initial results seem encouraging and support the interest of TNT strategy in achieving cCR and allowing for organ preservation.

These data must be interpreted in the context of a small population on patients. This study is ongoing, with further follow-up planned.

## Data Availability

Data are stored as de-identified participant data, which are available upon reasonable request from Luísa Leal-Costa.

## Ethical considerations

The study protocol was approved by the Hospital Beatriz Ângelo Ethics Committee (ref. CES/580_338). Written informed consent was obtained from all participants in this study.

## Acknowledgements

To all the elements that contributed to this work. Data are stored as de-identified participant data, which are available upon reasonable request from Luísa Leal-Costa.

